# Normative serum cortisol levels in second and third trimesters and their associated factors: A prospective cohort study from Sri Lanka

**DOI:** 10.1101/2025.03.27.25324819

**Authors:** Ishani Menike, Shashanka Rajapakse, Gayani Amarasinghe, Janith Warnasekara, Nuwan Darshana Wickramasinghe, Thilini Agampodi, Suneth Agampodi

**Affiliations:** District General Hospital Nuwaraeliya, 22200, Sri Lanka; Department of Physiology, Faculty of Medicine and Allied Sciences, Saliyapura, Anuradhapura, Sri Lanka; Graduate College of Cancer Science and Policy, National Cancer Center, 323 Ilsan-ro, Ilsandong-gu, Goyang-si Gyeonggi-do, 10408, Republic of Korea; Department of Community Medicine, Faculty of Medicine and Allied Sciences, Rajarata University of Sri Lanka, Saliyapura, 50008, Sri Lanka; International Vaccine Institute, SNU Research Park, 1 Gwanak-ro, Gwanak-gu, Seoul, 08826, Republic of Korea; Section of Infectious Diseases, Department of Internal Medicine, School of Medicine, Yale University, United States of America

**Author notes:** **Corresponding author** Suneth Agampodi. **Author contribution** Conceptualization: IM, SR, TA, SA Data collection: IM, GA, NDW, TA, SA Methodology: IM, SR, GA, NDW, TA, SA Investigation: IM, SR, GA, NDW, TA, SA Data curation: SA Formal analysis: IM, SR, JW, SA Writing-original draft: IM, SR Writing-review and editing: GA, JW, NDW, TA, SA Project administration: NDW, TA, SA Funding acquisition: TA, SA. **Data availability statement:** All deidentified data of the participants included in the current study of the Rajarata Pregnancy Cohort (RaPCO) is deposited under the doi 10.5281/zenodo.15074568 at the https://zenodo.org/records/15074568.

**Keywords:** Maternal Health, Maternal Stress, Cortisol, Rural Health, Pregnancy Cohort

## Abstract

**Objective:** To describe the normative serum cortisol levels during 25 to 29 weeks of POG and the association of maternal, psychological, and social factors on serum cortisol in the second and third trimesters in a cohort of pregnant women.

**Methods:** All eligible pregnant women registered in the maternal care program in Anuradhapura district, Sri Lanka from July to September 2019 were invited to the Rajarata Pregnancy Cohort (RaPCo). An interviewer-administered questionnaire-based symptom analysis and clinical assessment were conducted at baseline in the first trimester and at follow-up from 25 to 29 weeks POG. We assessed fasting early morning serum cortisol levels at the follow-up visit.

**Results:** The study sample included 1010 pregnant women with a mean age in years and POG in weeks at baseline of 28 (±6) and 10 (±3), respectively. The mean (SD, 97% percentile) serum cortisol level in all pregnant women was 10.93 (±3.83, 20.95) μg/dL, with no significant difference between singleton and twin pregnancies (*p*=0.138). None of the study participants had a cortisol level exceeding the upper limit of 42 μg/dL, and 464 (45.9%) had levels less than 10 μg/dL. Serum cortisol levels were higher in women with an advanced POG with a mean of 10.33 µg/dL (95%CI: 9.68–10.98) at 24 weeks POG and 12.23 µg/dL (95%CI: 11.15–13.32) at 29 weeks POG (*p*=0.049). Primi-gravidity (*p*=0.004), history of miscarriage (*p*=0.010), BMI categories (*p*=0.044), and POG (*p*=0.002) were independently associated with serum cortisol levels in robust regression. An EPDS score of more than 9 was not associated with serum cortisol (*p*=0.633).

**Conclusion:** The pregnant women in rural Sri Lanka reported a low mean serum cortisol level, which gradually increased with the POG. Significantly higher mean serum cortisol was associated with primi-gravidity, history of miscarriage, pre-pregnancy BMI, and POG at cortisol test but not psychological factors.

## Introduction

Maternal stress during pregnancy plays a critical role in maternal and fetal well-being. Stress during preconception and the antenatal period adversely affects maternal and fetal health through the dysregulation of the hypothalamic-pituitary axis [1]. Maternal stress increases the risk of maternal complications, including eclampsia, premature labor, and postpartum hemorrhage [2–4]. Recent evidence synthesis indicates maternal stress is implicated in intrauterine growth restriction, prematurity, and low birth weight [5]. It is also associated with developmental disorders and childhood physical and psychological disorders [6,7]. Thus, maternal stress during pregnancy and its causes should be explored in depth to ensure safe motherhood and the well-being of their offspring.

Currently, maternal stress is measured in two broad approaches: cortisol level and psychometric tools [8]. Different measures of cortisol, such as serum, urinary, hair, and diurnal cortisol, are currently used to assess stress [9]. Cortisol, the stress hormone, is considered a better objective indicator of physiological stress compared to the subjective evaluations conducted using psychometric tools [10–12]. Maternal cortisol, the final product of the hypothalamic-pituitary axis, is a key hormone in fetal development and maturation [13]. A recent study indicates that maternal serum total cortisol increased on average during pregnancy from 390 (±22) nmol/L in the 5^th^ week to 589 (±15) nmol/L in the 20^th^ week [14]. Current evidence suggests that high serum cortisol of more than 17.66 μg/dL (488 nmol/L) is significantly associated with developing postpartum depression [15]. However, studies from low socio-economic countries scarcely include measurement of cortisol levels in pregnancy [9].

The factors contributing to maternal stress are diverse and perceived maternal stress is associated with circumstantial factors such as marital status, unplanned pregnancy, societal factors such as support from extended family, factors such as teenage pregnancy, domestic abuse, psychological factors, and economic factors [16]. Furthermore, emotional lability and physiological changes in pregnancy such as weight gain, nausea, and insomnia also contribute to significant stress during pregnancy [17–19]. Cortisol is known to be associated with maternal depression, Y, and adverse birth outcomes [20]. This is further compounded in complicated pregnancies [21].

Sri Lanka is a lower-middle-income country with comparatively high maternal health indicators report high levels of antenatal anxiety, depression, and maternal suicides [22–24]. There are no studies reporting serum cortisol levels or predictors of cortisol in pregnant women of Sri Lanka. The current study investigates the normative data of serum cortisol levels in the second and third trimesters in a large population-based cohort and explores the association of maternal biological and psychosocial stressors on the serum cortisol level in the second trimester.

## Methodology

### Study design

We conducted a population-based prospective cohort study. The detailed methodology has been published [25].

### Study setting

This study was conducted in Anuradhapura, geographically the largest district in Sri Lanka. It is predominantly rural with mostly Sinhalese (91%) followed by Sri Lankan Moors (8.2%), Tamils (0.6%), and other minority groups (0.2%) [26]. The female literacy rate is 91.7% and 37.1% participate in economic activities in Anuradhapura, Sri Lanka [27]. Each year, over 15,000 expectant mothers enroll in public antenatal care services in Sri Lanka. These services are delivered through divisional units within the medical officer of health (MOH) areas. The district consists of 22 MOH areas, where all pregnant women are systematically registered by public health midwives (PHMM), either during field visits or at their initial antenatal appointment.

### Study population

All pregnant women residing in the Anuradhapura district, North Central Province, Sri Lanka, registered with the national pregnancy care program from 1^st^ July to 30^th^ September 2019 and with a period of amenorrhea of less than 12 weeks. The national maternal care program of Sri Lanka has 95% coverage [28].

### Study sample

Serum cortisol levels were available for 1290 cohort participants who attended the second or third-trimester follow-up. During the second visit, the period of gestation (POG) was recalculated from the dating scan. Out of them, 1010 pregnant women between 25 and 29 weeks of POG at the time of estimating serum cortisol level were selected for the current analysis to have an adequate number of women in each week of POG. Pregnant women were followed up from the first trimester to delivery.

### Procedure

At the baseline, all participants with 13 weeks of POG were included in the cohort study and underwent a detailed clinical interview, anthropometric assessments, and venipuncture for blood and serum investigations in addition to routine maternal care. Interviewer-based questionnaires were completed by medically trained data collectors to gather participants’ sociodemographic, medical, psychological, and social information. Anthropometric measurements included weight, height, waist circumference, and hip circumference to assess body mass index (BMI) and waist-to-hip ratio according to standard protocols.

During the follow-up visit, fasting early morning serum cortisol levels were obtained by a trained phlebotomist after adequate resting for each participant, obtaining blood in a single prick. The analytical work was conducted by an accredited external laboratory using a fully automated analyzer based on the electrochemiluminescence immune assay method. The validated and culturally-adopted Edinburgh Postnatal Depression Scale (EPDS) was used to assess antenatal depression during the first and second trimesters [29,30]. Furthermore, attempts or ideation of deliberate self-harm and/or suicide, and intimate partner violence and/or abuse were assessed in the second trimester.

### Statistical analysis

The distribution of cortisol levels from 25 to 29 weeks of POG was assessed for the whole study sample and singleton pregnancies. We considered a serum cortisol level of 10 μg/dL to 42 μg/dL as normal [31]. The serum cortisol level was described and compared based on the trimester, number of fetuses, gravidity, parity, maternal age, contraceptive failure, history of miscarriage, BMI category, waist-to-hip ratio, glycemic status, demographic factors, including maternal and paternal ethnicity, religion, and highest grade completed in school, and factors on mental health. The mean comparison of two groups and more than two groups were conducted with independent sample T-test and one-way ANOVA, respectively, with a significance (*p*) of 0.05. One-way ANOVA was conducted to compare the mean serum cortisol levels of women with a POG between 24 and 29 weeks with Levene’s test to test the heterogeneity of variances, and, if heterogeneous, Games-Howell post hoc comparisons were used. The association between total EPDS score and components (anhedonia, anxiety, and depression) in the first trimester and the mean serum cortisol level was explored with an independent sample T-test or one-way ANOVA [32]. Variables found to be significant (*p*<0.05) in the bivariate analysis were further evaluated using multivariable regression to identify independent associations with serum cortisol levels. In addition to these variables, marital status and maternal education were included in the model due to their potential confounding effects. Parity was excluded because of multicollinearity with gravidity. Given that serum cortisol was not normally distributed, the dependent variable was log-transformed to satisfy the assumptions of linear regression and improve model performance. After comparing model diagnostics, robust linear regression was selected over standard ordinary least-squares regression, as it provides more reliable estimates in the presence of outliers and non-constant variance in residuals.

### Ethical considerations

Ethical approval for the study was obtained from the Ethics Review Committee of the FMAS, RUSL (ERC/2019/). Informed written consent was obtained from the participant, and if the participant was less than 18 years of age, consent from parent(s) and/or guardian(s) and written assent from the participant was obtained before recruiting to the study.

## Results

### Characteristics of the study sample

The study sample included 1010 pregnant women between 15 to 45 years of age (Table 1). The mean (SD) age at conception was 27.6 (±5.7). There were 82 (8.1%) teenage pregnancies (age at conception less than 20 years), while most women belonged to the 25-29 age category (n=351, 34.8%). In this study sample, 23 (2.3%) were not married. Most women were in their first (n=317, 31.4%) or second pregnancy (n=316, 31.3%). Contraceptive failure was the reason for pregnancy in 26 (2.6%) of pregnancies. A history of miscarriage was reported by 189 (18.7%) women. Most participants belonged to the normal body mass index (n=361, 35.7%), and waist-to-hip ratio of more than 0.85 (n=349, 34.6%) categories.

**Table 1:**
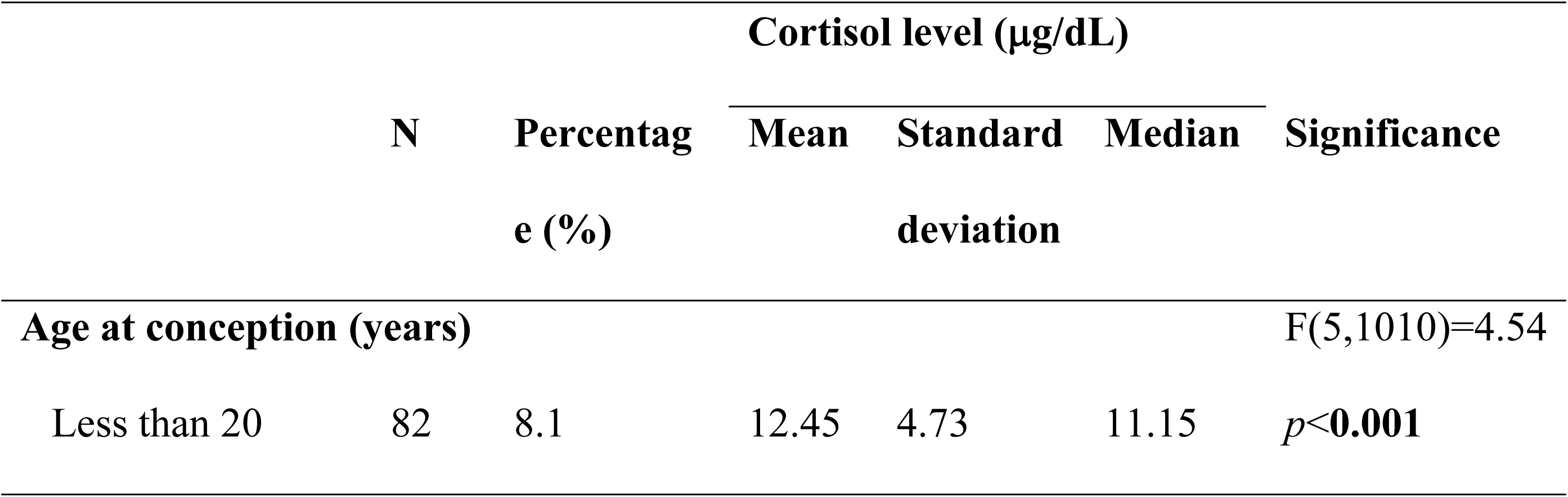
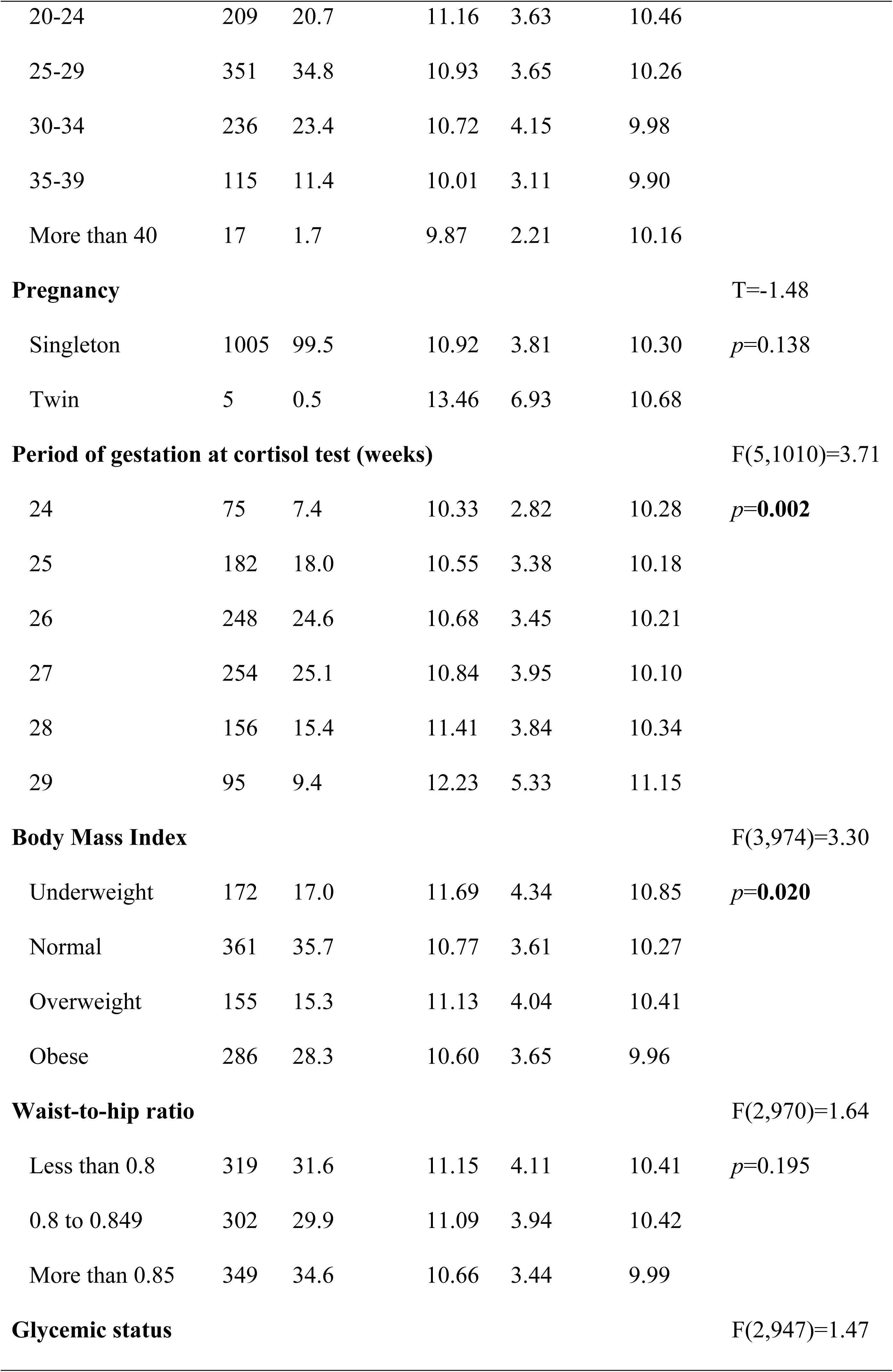
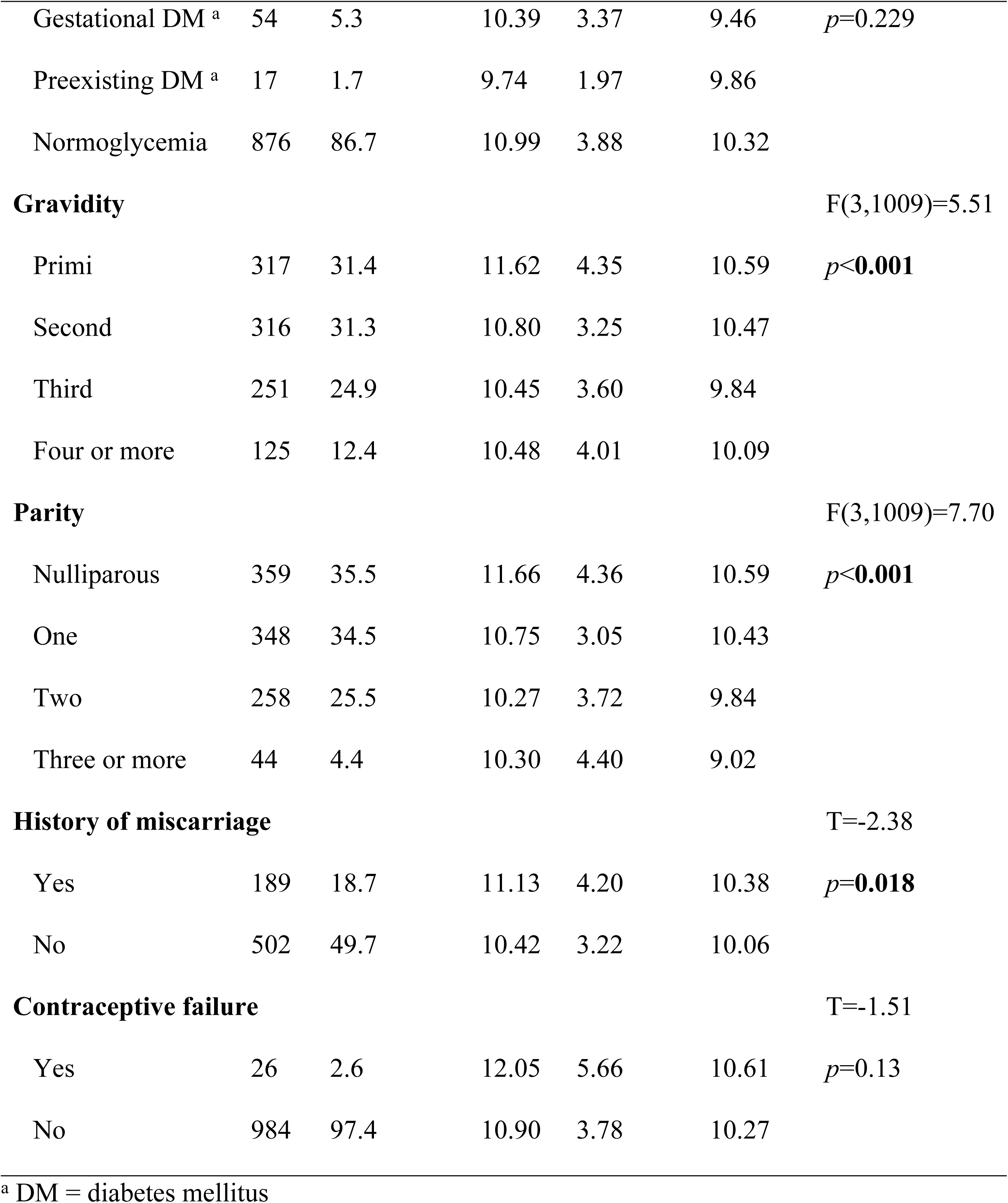
Biological factors affecting cortisol levels in the second and third trimester of pregnant women from Anuradhapura, Sri Lanka (N=1010).

| | N | Percentage (%) | Cortisol level ($\mu\text{g/dL}$) | | | Significance |
| --- | --- | --- | --- | --- | --- | --- |
|  |  |  | Mean | Standard deviation | Median |  |
| **Age at conception (years)** |  |  |  |  |  | F(5,1010)=4.54 |
| Less than 20 | 82 | 8.1 | 12.45 | 4.73 | 11.15 | *p*<0.001 |
| 20-24 | 209 | 20.7 | 11.16 | 3.63 | 10.46 |  |
| 25-29 | 351 | 34.8 | 10.93 | 3.65 | 10.26 |  |
| 30-34 | 236 | 23.4 | 10.72 | 4.15 | 9.98 |  |
| 35-39 | 115 | 11.4 | 10.01 | 3.11 | 9.90 |  |
| More than 40 | 17 | 1.7 | 9.87 | 2.21 | 10.16 |  |
| **Pregnancy** |  |  |  |  |  | T=-1.48 |
| Singleton | 1005 | 99.5 | 10.92 | 3.81 | 10.30 | *p*=0.138 |
| Twin | 5 | 0.5 | 13.46 | 6.93 | 10.68 |  |
| **Period of gestation at cortisol test (weeks)** |  |  |  |  |  | F(5,1010)=3.71 |
| 24 | 75 | 7.4 | 10.33 | 2.82 | 10.28 | *p*=**0.002** |
| 25 | 182 | 18.0 | 10.55 | 3.38 | 10.18 |  |
| 26 | 248 | 24.6 | 10.68 | 3.45 | 10.21 |  |
| 27 | 254 | 25.1 | 10.84 | 3.95 | 10.10 |  |
| 28 | 156 | 15.4 | 11.41 | 3.84 | 10.34 |  |
| 29 | 95 | 9.4 | 12.23 | 5.33 | 11.15 |  |
| **Body Mass Index** |  |  |  |  |  | F(3,974)=3.30 |
| Underweight | 172 | 17.0 | 11.69 | 4.34 | 10.85 | *p*=**0.020** |
| Normal | 361 | 35.7 | 10.77 | 3.61 | 10.27 |  |
| Overweight | 155 | 15.3 | 11.13 | 4.04 | 10.41 |  |
| Obese | 286 | 28.3 | 10.60 | 3.65 | 9.96 |  |
| **Waist-to-hip ratio** |  |  |  |  |  | F(2,970)=1.64 |
| Less than 0.8 | 319 | 31.6 | 11.15 | 4.11 | 10.41 | *p*=0.195 |
| 0.8 to 0.849 | 302 | 29.9 | 11.09 | 3.94 | 10.42 |  |
| More than 0.85 | 349 | 34.6 | 10.66 | 3.44 | 9.99 |  |
| **Glycemic status** |  |  |  |  |  | F(2,947)=1.47 |
| Gestational DM <sup>a</sup> | 54 | 5.3 | 10.39 | 3.37 | 9.46 | $p=0.229$ |
| Preexisting DM <sup>a</sup> | 17 | 1.7 | 9.74 | 1.97 | 9.86 |  |
| Normoglycemia | 876 | 86.7 | 10.99 | 3.88 | 10.32 |  |
| **Gravidity** | | | | | | $F(3,1009)=5.51$ |
| Primi | 317 | 31.4 | 11.62 | 4.35 | 10.59 | $p<0.001$ |
| Second | 316 | 31.3 | 10.80 | 3.25 | 10.47 |  |
| Third | 251 | 24.9 | 10.45 | 3.60 | 9.84 |  |
| Four or more | 125 | 12.4 | 10.48 | 4.01 | 10.09 |  |
| **Parity** | | | | | | $F(3,1009)=7.70$ |
| Nulliparous | 359 | 35.5 | 11.66 | 4.36 | 10.59 | $p<0.001$ |
| One | 348 | 34.5 | 10.75 | 3.05 | 10.43 |  |
| Two | 258 | 25.5 | 10.27 | 3.72 | 9.84 |  |
| Three or more | 44 | 4.4 | 10.30 | 4.40 | 9.02 |  |
| **History of miscarriage** | | | | | | $T=-2.38$ |
| Yes | 189 | 18.7 | 11.13 | 4.20 | 10.38 | $p=0.018$ |
| No | 502 | 49.7 | 10.42 | 3.22 | 10.06 |  |
| **Contraceptive failure** | | | | | | $T=-1.51$ |
| Yes | 26 | 2.6 | 12.05 | 5.66 | 10.61 | $p=0.13$ |
| No | 984 | 97.4 | 10.90 | 3.78 | 10.27 |  |

<sup>a</sup> DM = diabetes mellitus

### Normative serum cortisol data in 24 to 29 weeks of POG

The distribution of cortisol levels in this study was skewed right (Shapiro-Wilk =0.86, p<0.01). The mean (SD, 97.5% percentile) serum cortisol level of the study sample was 10.93 (±3.83, 20.95) μg/dL, and there was no significant difference (*p*=0.138) between the mean serum cortisol levels of singleton (10.92 ±3.81 μg/dL) and twin pregnancies (13.46±6.93 μg/dL) (Fig 1). None of the study participants had a cortisol level exceeding the upper limit of 42 μg/dL, and 464 (45.9%) had levels less than 10 μg/dL, with 236 (50.9%) in the second trimester and 228 (49.1%) in the third trimester. Serum cortisol levels gradually increased with the POG, with a mean of 10.33 µg/dL (95%CI: 9.68–10.98) at 24 weeks POG and 12.23 µg/dL (95%CI: 11.15–13.32) at 29 weeks POG (Fig 2). One-way ANOVA was conducted for those who are between 24 and 29 weeks, and it indicated a significant effect of POG on serum cortisol levels (F (5, 1010) =3.71, *p*=0.002) but with a small effect size (η²=0.02). Levene’s test was significant (*p*=0.002), indicating heterogeneity of variances, and, thus, Games-Howell post hoc comparisons were used. A significant difference was observed only between 24 weeks POG and 29 weeks POG (*p*=0.049), with no other comparisons reaching statistical significance.

**Fig 1:**
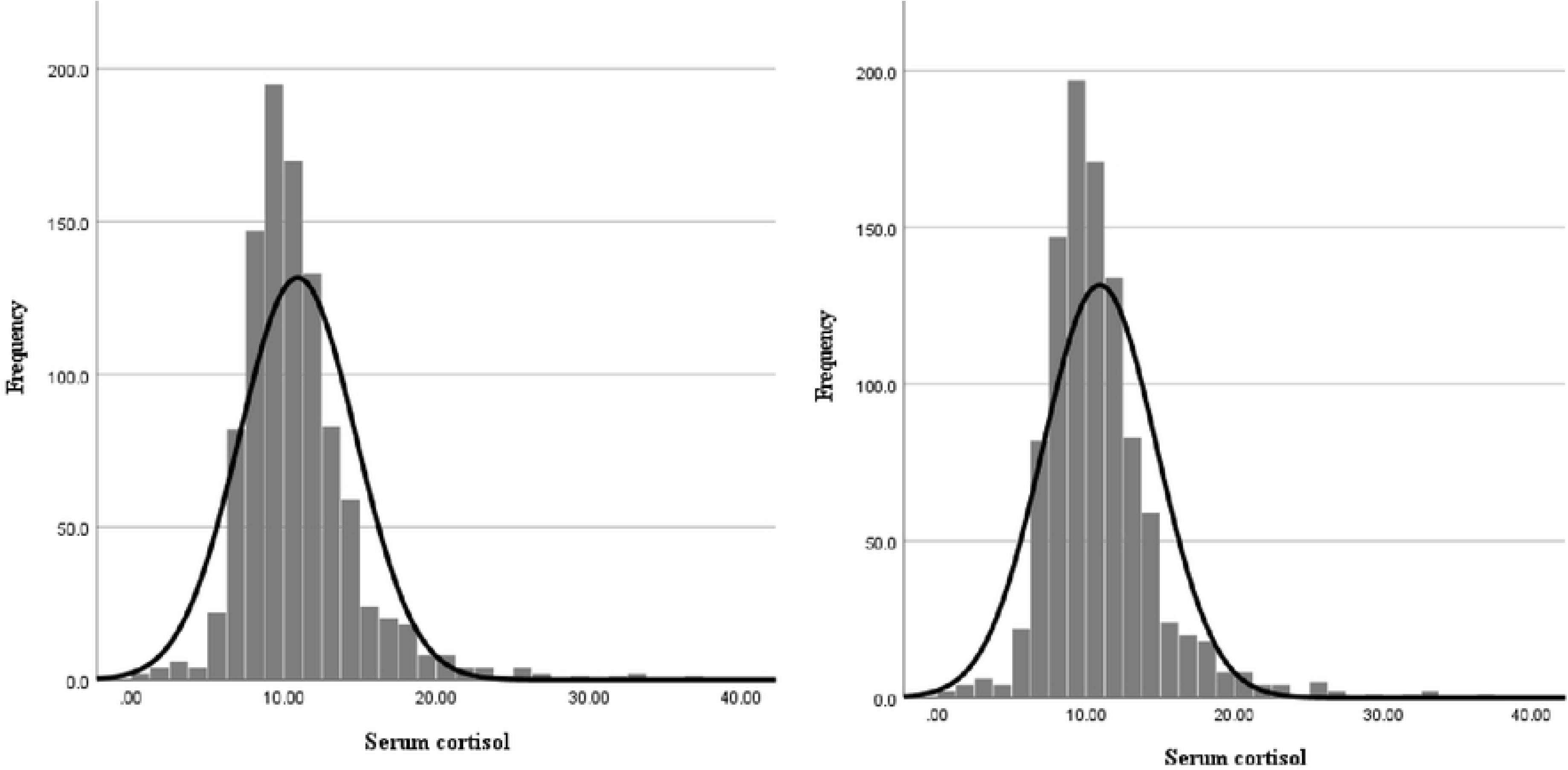
Distribution of cortisol in all pregnant women (left) and in singleton pregnancies (right).

**Fig 2:**
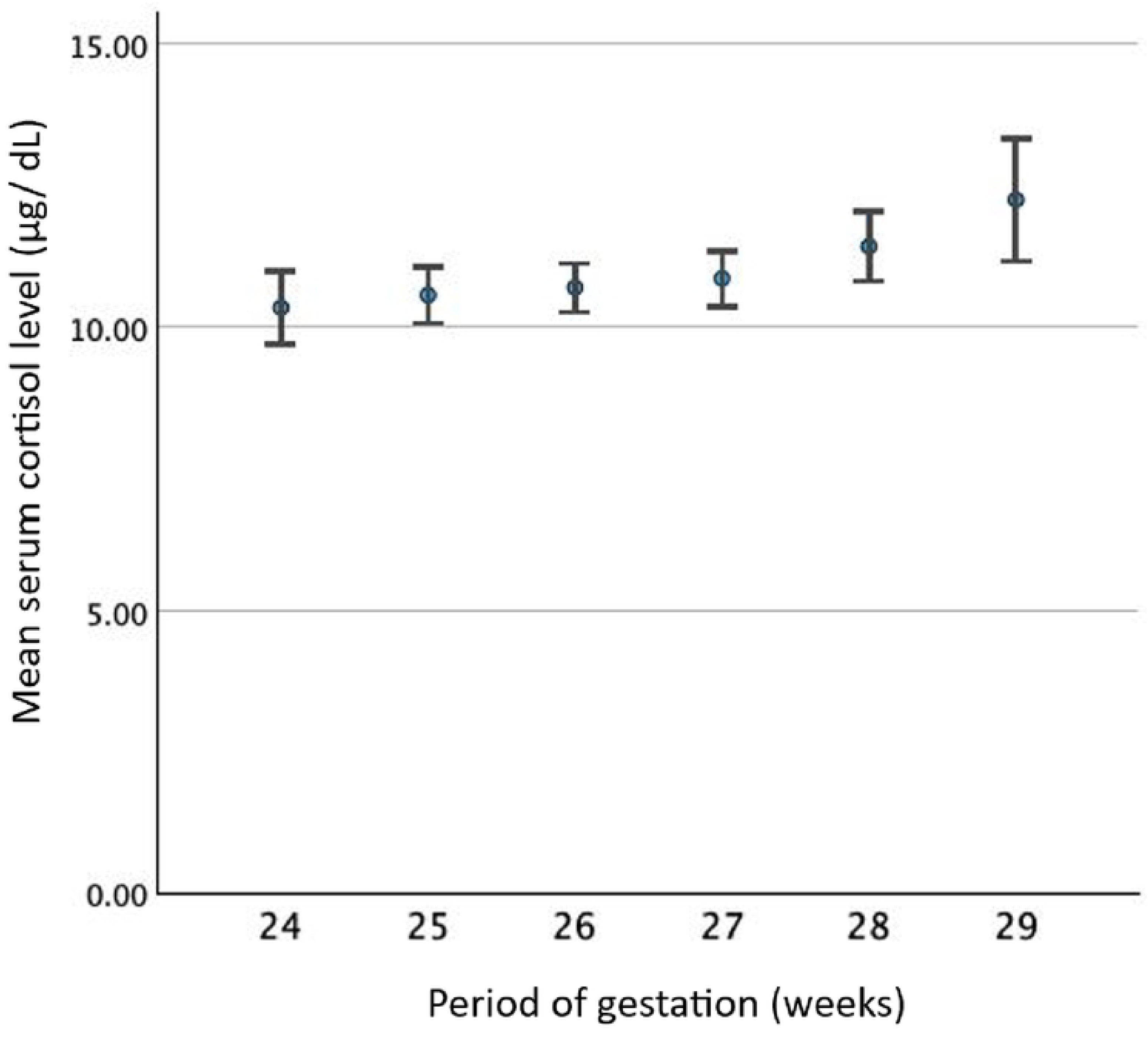
Distribution of mean serum cortisol level (μg/dL) by the period of gestation (POG) in weeks.

### Association of serum cortisol levels with biological and maternal factors

A significantly higher mean serum cortisol level was detected in primi-gravida (one-way ANOVA F (3, 1009) =5.51, *p*<0.001), nulliparous women (one-way ANOVA F (3, 1009) =7.70, *p*<0.001), teenage pregnancies (T=3.78 *p*<0.001) and women with a history of miscarriage (T=-2.38, *p*=0.018) compared to their respective counterparts (Fig 3). There was a significant difference between the mean serum cortisol levels of underweight, normal weight, overweight, and obese individuals (one-way ANOVA F (3, 974) =3.30, *p*=0.020). However, there was no significant difference in the mean serum cortisol levels between the groups based on the waist-to-hip ratio (one-way ANOVA F (2, 970) =1.64, *p*=0.195) or the glycemic status (gestational diabetes mellitus, pre-existing diabetes mellitus, and normoglycemia) (one-way ANOVA F (2, 947) =1.47, *p*=0.229). Maternal ethnicity (one-way ANOVA F (2, 1000) =0.414, *p*=0.661), religion (one-way ANOVA F (2, 1010) =0.827, *p*=0.438), highest grade completed in school (one-way ANOVA F (2, 1003) =0.073, *p*=0.930) or paternal ethnicity (one-way ANOVA F (2, 1007) =0.300, *p*=0.741), religion (one-way ANOVA F (2, 1008) =0.614, *p*=0.541), highest grade completed in school (one-way ANOVA F (2, 998) =1.057, *p*=0.350) were associated with a higher mean serum cortisol level.

**Fig 3:**
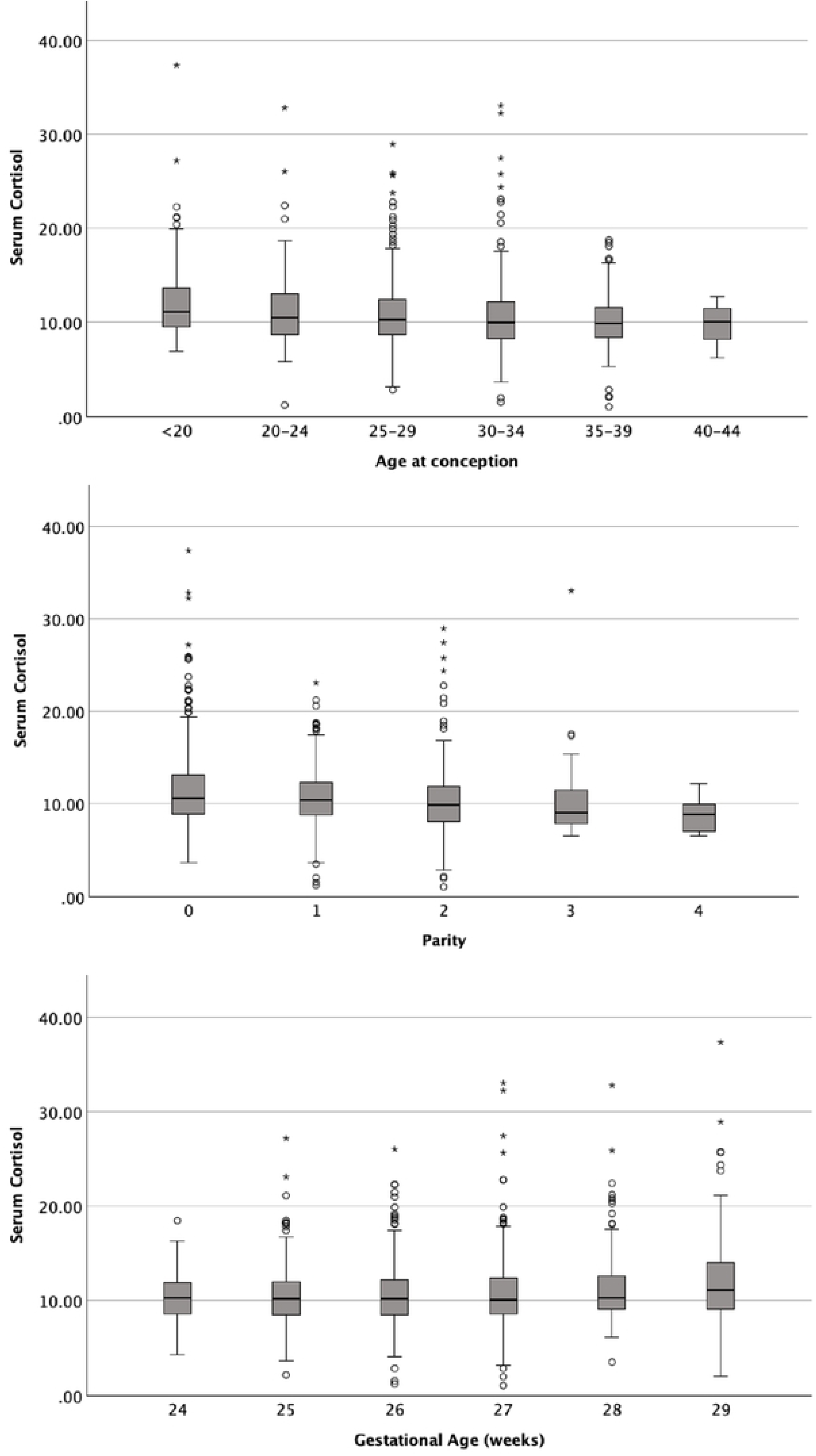
Distribution of serum cortisol level by the age at conception in years (top), parity (middle), and gestational age in completed weeks (bottom).

### Association of serum cortisol levels with sociodemographic and psychological factors

An EPDS score of more than 9 was not associated with serum cortisol (*p*=0.633). The first-trimester EPDS scores of anhedonia (one-way ANOVA F(641,963)=1.01, *p*=0.444), anxiety (one-way ANOVA F(641,963)=0.96, *p*=0.655), or depression (due to low numbers with each score, participants with a score of 10 or more were amalgamated into one group) (one-way ANOVA F(641,963)=1.13, *p*=0.103) in the first trimester did not predict mean serum cortisol in the second and third trimester (Table 2). Similarly, suicidal attempts (*p*=0.185), suicidal ideation (*p*=0.572), and attempts at deliberate self-harm (*p*=0.631) were not associated with serum cortisol levels. The serum cortisol level was not associated with the EPDS score in the second trimester (one-way ANOVA F (599, 872) =0.96, *p*=0.663).

**Table 2:**
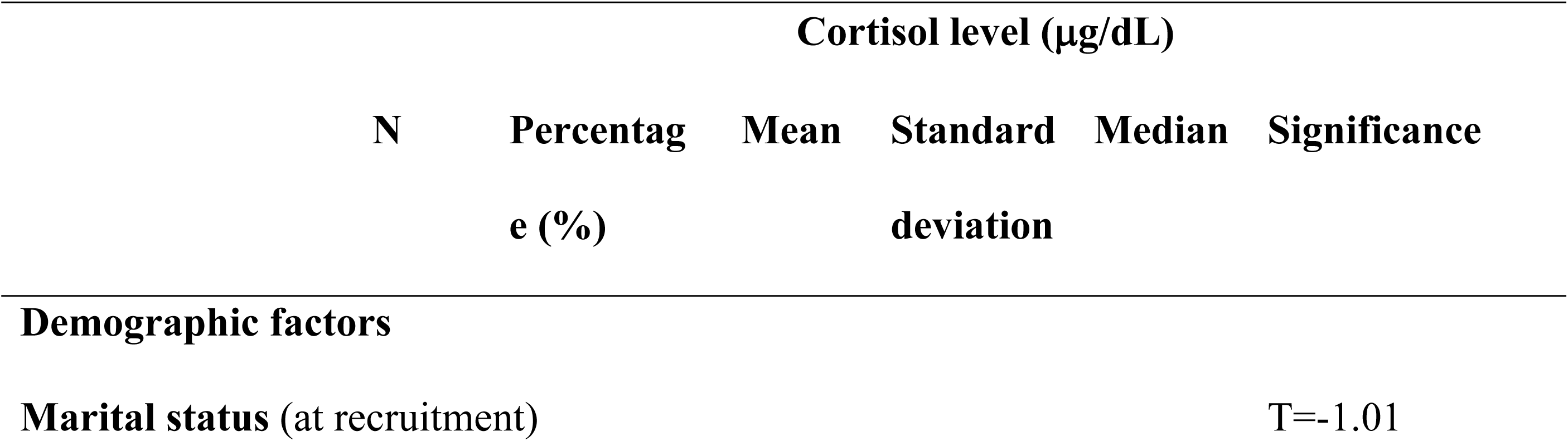
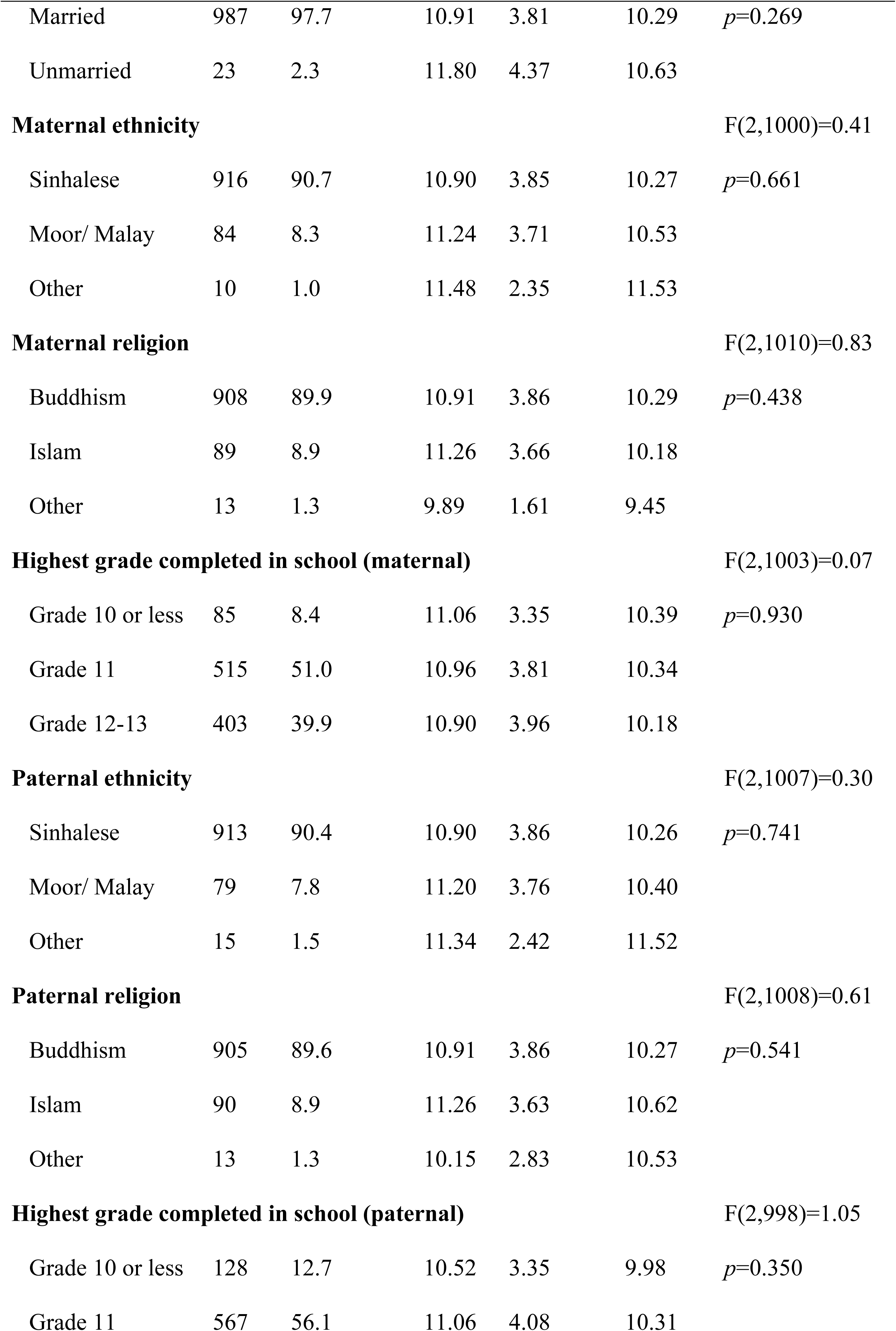
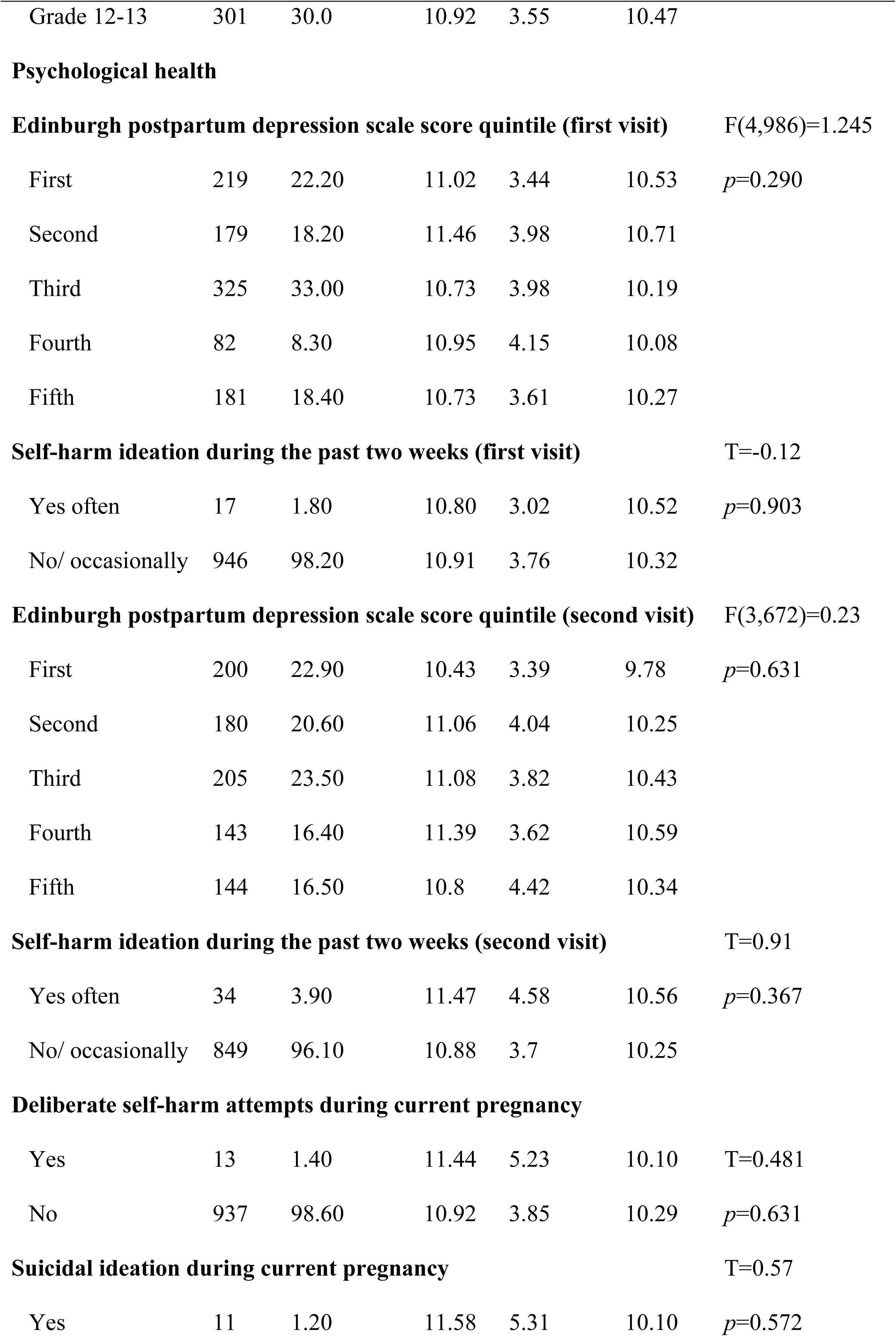
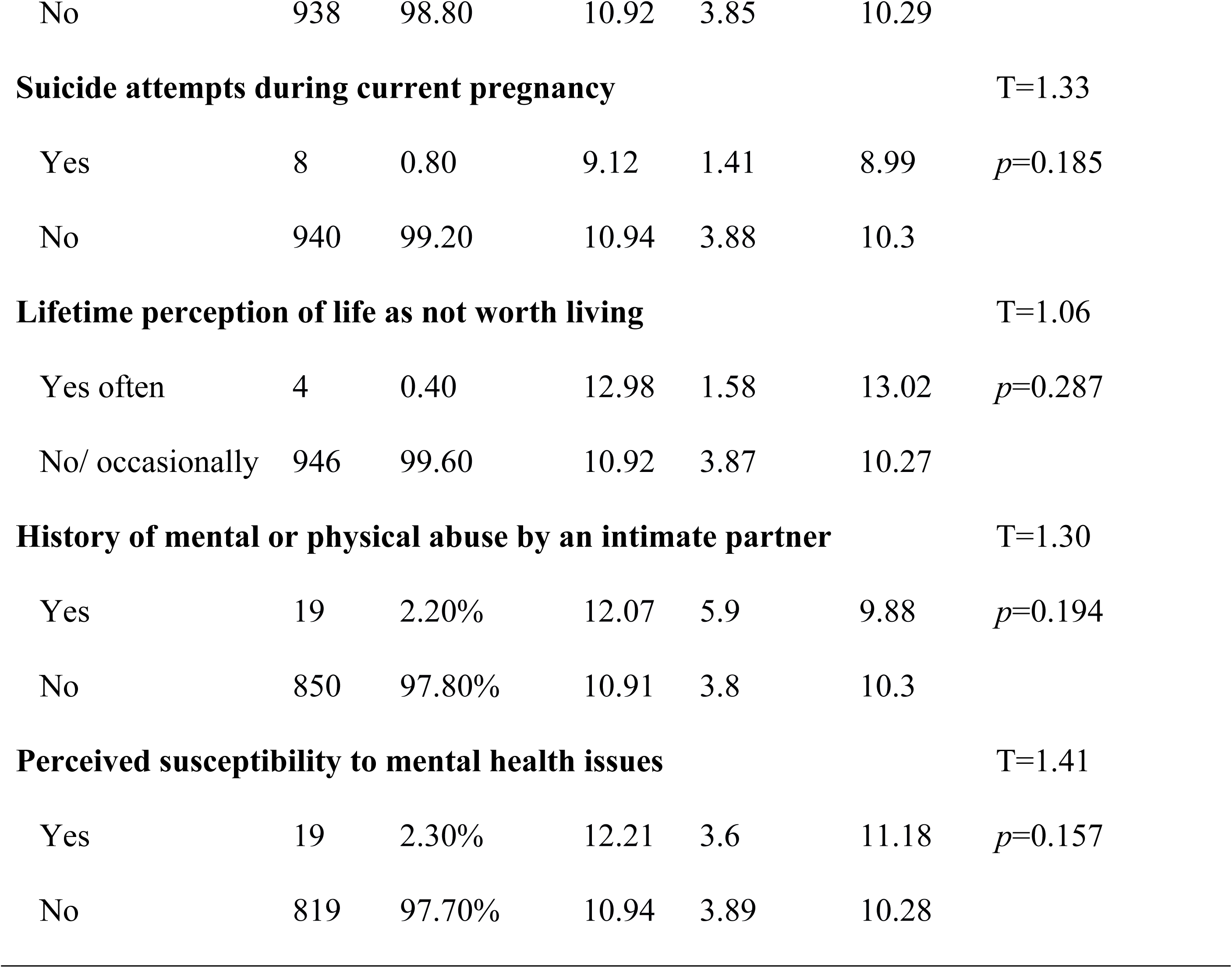
Sociodemographic factors and psychological factors affecting cortisol levels in the second and third trimesters in pregnant women from Anuradhapura, Sri Lanka (N=1010).

### Independent predictors of serum cortisol level

The robust regression analysis showed that a history of miscarriages (ß=0.081, 95%CI 0.020 to 0.142, *p*=0.010) and POG at cortisol test (ß=0.015, 95%CI 0.006 to 0.025, *p*=0.002) were positively associated with serum cortisol levels, whereas BMI (ß= −0.004, 95%CI −0.009 to - 0.001, *p*=0.044) and gravidity (ß= −0.044, 95%CI −0.074 to −0.014, *p*=0.004) were negatively associated (Table 3). These associations remained significant after adjusting for potential confounders.

**Table 3:** Robust Regression to determine independent predictors of serum cortisol level.

| Variable | df | Coefficient | 95%CI |  | Significance<br>(*p*-value) |
| --- | --- | --- | --- | --- | --- |
|  |  |  | lower | upper |  |
| **Constant** | 822 | 2.164 | 1.787 | 2.540 | <0.001 |
| **Age** | 822 | -0.002 | -0.006 | 0.003 | 0.505 |
| **Education level** | 822 | -0.004 | -0.018 | 0.009 | 0.524 |
| **Marital status** | 822 | 0.036 | -0.101 | 0.173 | 0.605 |
| **Gravidity** | 822 | -0.044 | -0.074 | -0.014 | 0.004 |
| **History of miscarriages** | 822 | 0.081 | 0.020 | 0.142 | 0.010 |
| **Body-mass-index** | 822 | -0.004 | -0.009 | -0.001 | 0.044 |
| **Period of gestation at cortisol test** | 822 | 0.015 | 0.006 | 0.025 | 0.002 |

## Discussion

We report normative data for serum cortisol levels in pregnancy and examine the factors associated with maternal serum cortisol in a large population-based cohort from rural Sri Lanka for the first time in Sri Lanka. The mean serum cortisol level in the study sample was close to the lower limit with none of the study participants exceeding the upper limit of the standard levels [31]. There was no significant difference between singleton and twin pregnancies. Serum cortisol levels in participants with 29-week POG were higher than in women with 24-week POG. The robust regression analysis showed that a history of miscarriages and POG at the cortisol test were positively associated with serum cortisol levels, whereas BMI and gravidity were negatively associated. We did not observe an association of serum cortisol with psychosocial factors.

### Normative levels

The current study includes all pregnant women enrolled in the maternal care program from the Anuradhapura district, Sri Lanka validating the normative data presented. In this study, the mean serum cortisol level was low and the upper limit of cortisol level was not exceeded in any participant. However, this result should be interpreted with caution as chronic stress may not result in elevated serum cortisol levels [33]. The mean cortisol levels reported from Southern Brazil in the first, second, and third trimesters were 28.5 μg/dl (95%CI: 26.1 to 31.2), 42.6 μg/dl (95%CI: 41.0 to 44.2), and 54.4 μg/dl (95%CI: 51.4 to 57.7), respectively with an increase of 2.0% (95%CI 1.01 to 1.02, *p*<0.001) each week [34]. Normal levels reported in Melbourne, Australia in the first, second, and third trimesters were 20.92 ±1.09 μg/dL, 31.84 ± 1.01 μg/dL, and 37.81 ± 1.49 μg/dL, respectively. However, further prospective cohort studies should be conducted in different geographies and populations to corroborate these findings. The non-significant difference between singleton and twin pregnancies in the current study is probably attributable to the low number of twin pregnancies, which resulted in a large standard deviation despite the apparent difference in the mean. The increase in cortisol levels with POG follows the known pattern of increasing from the first to the third trimester and during labor [12,14,35].

### Maternal factors

We report significantly higher serum cortisol levels in primi, nulliparous, and teenage pregnant women and women with past miscarriages compared to their counterparts. Robust regression was used to account for the presence of outliers and deviations from the assumptions of standard linear regression. This approach enhanced the reliability of our findings, especially given the skewed nature of cortisol data and biological variability among participants. It showed similar results: a positive association between POG and a history of miscarriage, and an inverse relationship with BMI and gravidity. The negative association of gravidity with serum cortisol levels may reflect physiological or psychological adaptation in women with previous pregnancies [36]. It is possible that women who have experienced multiple pregnancies develop better-coping mechanisms or reduced hypothalamic-pituitary-adrenal (HPA) axis reactivity over time. In contrast, the positive association between a history of miscarriages and cortisol levels even in the second and third trimesters may indicate increased stress or anxiety in subsequent pregnancies, which has not been adequately explored. The negative association of gravidity with serum cortisol levels may reflect physiological or psychological adaptation in women with previous pregnancies [36]. It is possible that women who have experienced multiple pregnancies develop better coping mechanisms or reduced hypothalamic-pituitary-adrenal (HPA) axis reactivity over time. In contrast, the positive association between a history of miscarriages and cortisol levels even in the second and third trimesters may indicate increased stress or anxiety in subsequent pregnancies, which has not been adequately explored. Similar to the current study, there was a significant difference between the mean serum cortisol levels of underweight, normal-weight, overweight, and obese individuals in other studies [37]. Several studies have shown that women with pre-pregnancy obesity have a lower level of salivary [38] and serum cortisol compared to lean women without an increase in urinary glucocorticoid clearance suggestive of decreased maternal hypothalamic-pituitary axis activity [39].

Known stressors in early pregnancy, such as marital status and contraceptive failure, were not significantly associated with serum cortisol levels in this study. However, contrary results similar to the current study were reported from a prospective cohort study from Bangalore, where a significant difference in cortisol levels was found for maternal age and gravidity but no other pregnancy-related stressors such as history of abortion, social support, and spousal alcohol consumption or smoking [15].

### Serum cortisol and psychosocial factors

Current study results did not show an association between the serum cortisol level in 24 to 29 weeks of POG with the EPDS score or other psychosocial factors. However, this may be attributable to the extensive supportive program, which included targeted interventions such as counseling, health education, financial and social support, and psychiatric referral as appropriate, conducted for the participants with high EPDS or psychological risk factors in the first-trimester screening [40]. Previous studies have shown mixed results on the association between subjective measures of depression or stress and serum cortisol levels [41,42]. However, current evidence favors maternal cortisol levels over subjective measures as a predictor of offspring outcomes [11]. Therefore, it is recommended to assess maternal cortisol in future studies and explore the public health implications of high maternal serum cortisol.

### Strengths and limitations

This is a prospective cohort study with a large sample size with only 3 other studies having a larger sample size, which were mainly from the developed countries. Furthermore, this is a population-based cohort study including all pregnant women residing in the Anuradhapura district, North Central Province, Sri Lanka, and registered with the national maternal care program from 1^st^ July to 30^th^ September 2019 and with a POG of less than 13 weeks. The national maternal care program has 95% antenatal coverage. Therefore, this study sample includes a representative population. A structured questionnaire administered by trained medical care personnel was used to evaluate psychological stressors. Furthermore, physical measurements were conducted by trained medical personnel. The fasting early-morning serum cortisol levels were evaluated in all participants after an adequate test by a trained phlebotomist and evaluated on the same day at an accredited laboratory. The current study did not evaluate hair or salivary cortisol levels for corroboration with serum cortisol levels due to budgetary concerns and the lack of accredited laboratory facilities.

### Conclusion

The mean serum cortisol level in all pregnant women was 10.93(±3.83) μg/dL, with no significant difference between singleton and twin pregnancies. Serum cortisol levels gradually increased with the POG, with a significant difference observed between POG 24 and POG 29 weeks. A history of miscarriages and POG at cortisol test were positively associated with serum cortisol levels, whereas BMI and were negatively associated. A high EPDS score or other psychological stressors were not associated with the serum cortisol level.

## Data Availability

All deidentified data of the participants included in the current study of the Rajarata Pregnancy Cohort (RaPCO) is deposited under the doi 10.5281/zenodo.15074568 at the https://zenodo.org/records/15074568.

## Acknowledgments

The author Shashanka Rajapakse expresses their appreciation for the support from the International Cooperation & Education Program (NCCRI·NCCI 52210-52211, 2024) of the National Cancer Center, Korea.

